# Synthetic modified vaccinia Ankara vaccines confer potent monkeypox immunity in non-human primates and healthy adults

**DOI:** 10.1101/2022.07.26.22277958

**Authors:** Flavia Chiuppesi, John A. Zaia, Sandra Ortega Francisco, Minh Ly, Felix Wussow, Don J. Diamond

## Abstract

The recent outbreak of monkeypox (MPXV) outside its endemic boundaries has attracted global attention and prompted world leaders to reserve millions of doses of the only approved third-generation smallpox/MPXV vaccine, Jynneos, which is based on the highly attenuated modified vaccinia Ankara (MVA) vector. We previously developed COH04S1, a multiantigen SARS-CoV-2 vaccine built on a synthetic MVA (sMVA) platform. COH04S1 was extensively tested for efficacy and immunogenicity in animal models, including non-human primates (NHP), and was found to be safe and to induce SARS-CoV-2-specific immunity in a Phase 1 clinical trial in healthy adults. Here we demonstrate that one or two vaccinations of NHP with either COH04S1 or sMVA elicit robust othopoxvirus-specific binding and neutralizing antibody responses. Furthermore, healthy adults vaccinated with COH04S1 at different dose levels develop robust othopoxvirus-specific humoral and cellular immune responses that are durable for over six months post-vaccination. Importantly, both COH04S1 and sMVA vaccinations induce elevated and sustained antibody responses to MPXV-proteins that are major targets of protective neutralizing antibodies. These results demonstrate that COH04S1 and sMVA are valuable vaccine candidates to stimulate robust orthopox/MPXV-specific humoral and cellular immunity.

## Introduction

The unprecedented 2022 monkeypox (MPXV) outbreak outside of its endemic boundaries has worried health officials and renewed interest in testing and stockpiling of a safe and effective smallpox/MPXV vaccine^1^. Smallpox (or *variola major*), a poxvirus with a case-fatality rate of up to 30%, was declared eradicated in 1980 after a very successful global vaccination campaign^2^. MPXV belongs to the same *orthopoxvirus* genus as smallpox and exists in two clades. The Central African clade has a high mortality rate of 10%, while infection with the West African clade results in about 1% case-fatality rate^3^. MPXV is endemic in Central African countries, where it causes more than 1,000 cases annually^3-5^. The recent outbreak with epicenter in Europe caused by the less severe West African clade has resulted in more than 30,000 cases (as of August 8^th^, 2022) with nine reported deaths globally. Recently reported cases in children and healthcare workers has prompted the WHO to declare the MPXV outbreak a global health emergency. Similarly, the federal and many state governments of the USA also declared MPXV a public health emergency on August 4^th^, 2022.

Replication-competent vaccinia virus strains of different origin were used worldwide for the smallpox vaccination campaign, which ended in 1972^6^. In the US, vaccinia strain Dryvax grown on calf skin formed the first-generation smallpox vaccine and was later substituted by ACAM2000, which was plaque purified from Dryvax and produced using modern cell culture technology. While ACAM2000 was highly immunogenic, it was associated with a high risk of myocarditis/pericarditis (1 in 175 naïve adults), and this risk also extended to close contacts of vaccinated subjects, posing a threat, albeit low, for children, pregnant women, and immunocompromised individuals^6^. For this reason, use of ACAM2000 has been licensed with a medication guide and its use restricted to designated U.S. military personnel and laboratory researchers working with certain poxviruses.

Modified vaccinia Ankara (MVA) is a highly attenuated, replication-defective orthopoxvirus that was derived from its parental strain chorioallantois vaccinia virus Ankara by over 500 passages on chicken embryo fibroblasts^7^. MVA was developed as a third-generation smallpox vaccine and has been safely administered at the end of the smallpox eradication campaign to more than 120,000 individuals, including children with immunodeficiencies and HIV infected individuals^8^. Recently, Bavarian Nordic’s proprietary MVA strain MVA-BN has been approved by the FDA as a smallpox and MPXV vaccine under the name of Jynneos based on a phase 3 non-inferiority trial comparing one dose of ACAM2000 with two intramuscular doses of Jynneos^9^ as well as nonhuman primate (NHP) studies using a lethal MPXV virus challenge^10^. Consequently, Jynneos has been added to the Strategic National Stockpile (SNS) as a safer alternative to ACAM2000 that could be administered to the broader population. Due to the recent MPXV outbreak, Jynneos is now offered as a prophylactic vaccine in at-risk subjects and for ring vaccinations in possible contacts of infected individuals. Given its robust safety and immunogenicity profile, MVA has also been extensively used as a viral vector for delivery of heterologous antigens and tested as a vaccine against infectious diseases and cancer^8,11-13^.

We developed a fully synthetic MVA (sMVA) platform based on the genome sequence published by Antione *et al*.^14^ for reconstituting virus that is virtually identical to wild-type MVA in terms of replication properties, host cell tropism, and immunogenicity^15^. Using this platform, which allows rapid generation of sMVA recombinants encoding multiple transgenes, we developed COH04S1, a multiantigen sMVA-based COVID-19 vaccine encoding for spike (S) and nucleocapsid (N) antigens. COH04S1 has been extensively tested in small and large animal models, demonstrating robust immunogenicity and protective efficacy against SARS-CoV-2 and its variants through intramuscular and intranasal routes of vaccination^15-17^. Additionally, COH04S1 has been tested in a phase I, randomized, placebo-controlled clinical trial in healthy adults, showing a remarkable safety profile and resulting in the induction of robust and durable humoral and cellular responses to both vaccine antigens^18,19^. Currently, COH04S1 is being evaluated in two phase 2 clinical trials in healthy adults and immunocompromised patients (NCT04639466, NCT04977024).

Given the recent approval of MVA as a vaccine against smallpox and MPXV, we evaluated whether COH04S1- or sMVA-vaccinated NHP^16^ and COH04S1-vaccinated healthy volunteers^18^ mount orthopoxviral-specific immunity. We found that NHP vaccinated with either COH04S1 or sMVA, and COH04S1-vaccinated healthy adults develop robust orthopoxviral-specific humoral and cellular immune responses, including antibodies to MPXV-specific proteins that are major targets of protective neutralizing antibody (NAb) responses, indicating that COH04S1 and sMVA represent unique vaccine candidates to control the unforeseen global MPXV outbreak.

## Results

### COH04S1 and sMVA induce robust orthopoxviral immunity in non-human primates

We retrospectively evaluated orthopoxviral-specific humoral responses in NHP vaccinated with one or two doses of COH04S1 or empty sMVA control vector. NHP were either vaccinated once with a higher dose (5×10^8^ plaque forming units [pfu]), or they were vaccinated twice with half of the vaccine dose (2.5×10^8^ pfu/dose). Mock-vaccinated NHP were used as controls. MVA-specific IgG were evaluated in NHP serum by ELISA. With the exception of one NHP with low-level MVA-specific IgG, no orthopoxviral-specific pre-existing immunity was observed in pre-immune samples. In contrast, at one-month after the first vaccination all NHP vaccinated with either COH04S1 or sMVA developed robust MVA-specific IgG independently of the used vaccine dose. NHP vaccinated once at a higher vaccine dose tended to have higher median MVA-specific IgG endpoint titers than NHP vaccinated once at a lower vaccine dose. In contrast, in NHP receiving a second lower vaccine dose, MVA-specific IgG were boosted in both COH04S1- and sMVA-vaccinated animals and tended to exceed those induced in NHP receiving a single shot at higher vaccine dose. No differences in IgG endpoint titers were observed between sMVA- and COH04S1-vaccinated NHP (Figs. 1A-B and S1).

**Figure 1.**
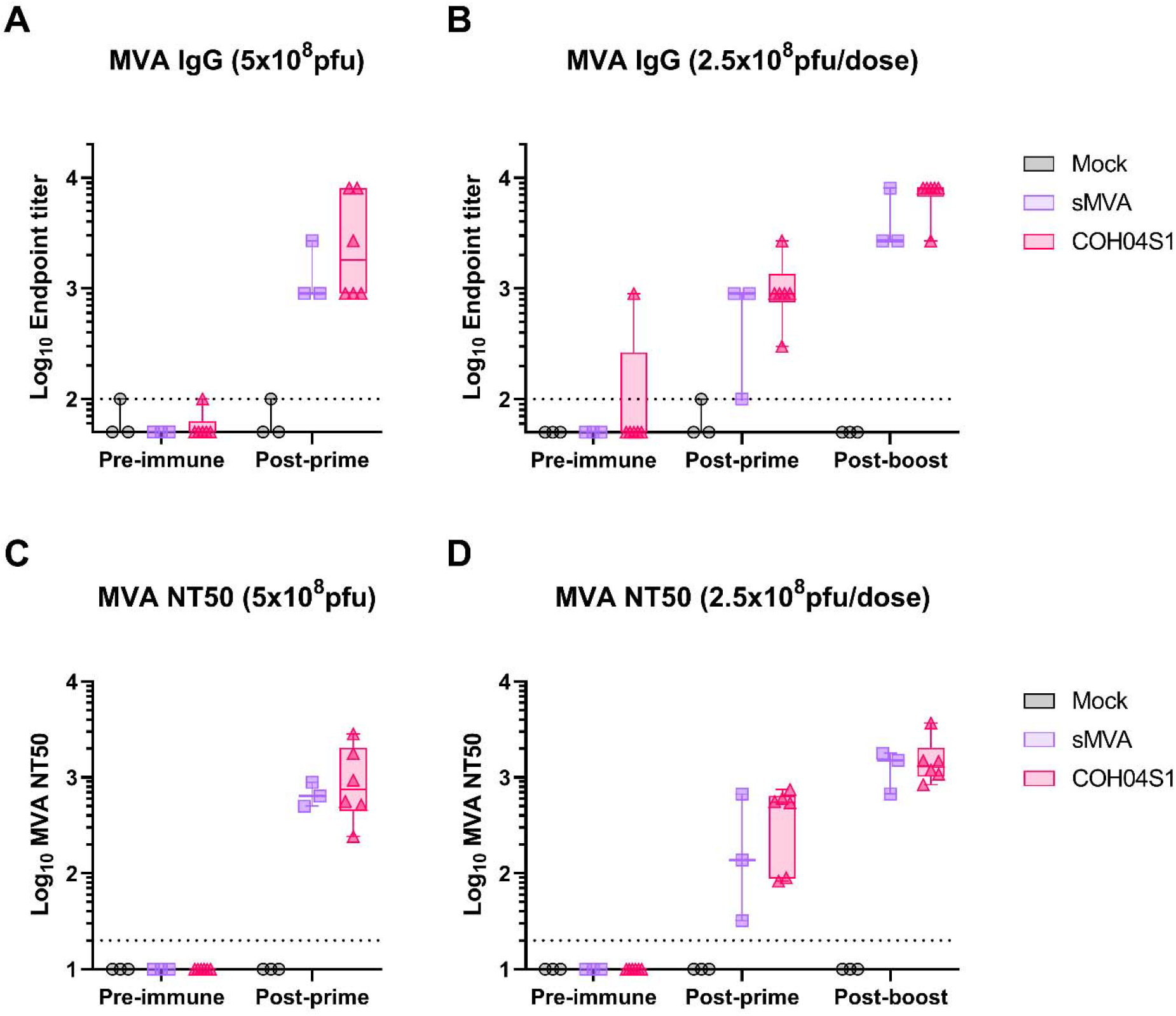
MVA-specific humoral responses in sMVA- and COH04S1-vaccinated NHP. NHP were vaccinated once with 5×10^8^ pfu (A, C) or two-times vaccinated with 2.5×10^8^ pfu (B, D) of sMVA (n=3) or COH04S1 (n=6). Mock-vaccinated NHP were used as controls (n=3). **A-B**. MVA-specific IgG endpoint titers were measured by ELISA at baseline, one month after the first dose, and one month after the second dose (in B). **C-D**. MVA-specific NAb titers. NAb specific for MVA were measured by microneutralization assay at baseline, one month after the first dose, and one month after the second dose (in D). Dotted lines represent the lower limit of detection of the assay. Box plots extend from the 25^th^ to the 75^th^ percentiles, median values are shown as a line, whiskers extend from minimum to maximum values.

MVA-specific NAb were measured on epithelial cells using a high-throughput microneutralization assay. Both one- and two-dose vaccination regimens elicited potent MVA-specific NAb titers. At one month after the first vaccination NHP vaccinated with COH04S1 or sMVA at lower vaccine dose developed MVA-specific NAb titers (NT50) that ranged from 30 to 750, while higher MVA-specific NAb titers ranging from 240 to 2,840 were induced in NHP vaccinated once at higher vaccine dose. In NHP vaccinated with a second injection of low vaccine dose, NAb were boosted in both COH04S1 and sMVA-vaccinated animals to NAb titers ranging from 680 to 3,690. Similar NAb titers were measured between COH04S1- and sMVA-vaccinated NHP (Figs. 1C-D and S1). These results demonstrate that both sMVA-based vaccine COH04S1 and sMVA itself promote robust induction of MVA-specific humoral responses after single vaccine dose, while a second dose can increase the magnitude of the vaccine-elicited antibody responses.

### COH04S1-vaccinated subjects develop robust orthopoxviral-specific binding antibodies

Next, we retrospectively evaluated orthopoxviral-specific humoral and cellular immune responses for up to six months after vaccination with COH04S1 in a subgroup of 20 volunteers enrolled in a phase 1 clinical trial aimed at testing safety and immunogenicity of COH04S1 at different dose levels (DL) (NCT04639466)^18,19^. Subjects were prime-boost vaccinated with low-dose (DL1, 1×10^7^ pfu), medium-dose (DL2, 1×10^8^ pfu), or high-dose (DL3, 2.5×10^8^ pfu) of vaccine. Of the 20 subjects vaccinated with COH04S1, 15 (5 subjects/group) received two DL1, DL2, or DL3 vaccinations 28 days apart, and 5 received two DL1 vaccinations 56 days apart with a placebo dose at day 28 (DL1/placebo/DL1). Four placebo-vaccinated subjects enrolled in the same trial were included as controls. Subjects were not required to provide their smallpox vaccination status, and poxvirus serostatus at enrollment was not evaluated. However, an exclusion criterion was any poxvirus-vaccination in the six months before enrolling in the trial. Summary of study subjects, vaccination schedule and age at enrollment is presented on Table S1.

MVA-specific IgG binding antibodies in serum of COH04S1 vaccinated subjects were measured against whole MVA virions by ELISA. Low binding (O.D.<0.4 nm) at low serum dilution (1:150) was measured at baseline in most subjects (Figure S2). In contrast to all placebo control volunteers, all subjects vaccinated with COH04S1 showed an increase in MVA-specific IgG titers post-vaccination regardless of the dose vaccination regimen (Figure 2), demonstrating potent vaccine-elicited orthopoxviral-specific humoral immunity. Elevated MVA-specific IgG titers were measured following prime vaccination at all dose levels, although MVA IgG titers in DL2 and DL3 subjects tended to be higher than those in DL1 subjects, indicating a dose dependent response (Figure 2A-B). While DL1 cohorts had a seroconversion rate of 30-60% following prime vaccination, DL2 and DL3 subjects showed 100% seroconversion after the first dose (Figure 2C). MVA-specific IgG titers further increased in all vaccine cohorts following the second dose resulting in similar responses in DL1 and DL2/3 subjects and 100% seroconversion in all vaccine cohorts. MVA-specific IgG titers slowly declined over five-months post vaccination in all vaccine cohorts, but they remained at elevated levels over baseline in all subjects independent of the dose immunization regimen, except for one subject in the DL1 cohort. Notably, two subjects in the DL1 and DL3 cohorts - one subject born in 1971 and one subject born in 1986 - had high IgG endpoint titers of 4,050 and 1,350 before vaccination, possibly indicating pre-existing orthopoxviral immunity (Figure S2). These two subjects showed particularly elevated MVA-IgG titers after only one vaccine dose and their MVA IgG titers remained stable over six months post vaccination. These results demonstrate that healthy volunteers vaccinated with COH04S1 at different dose levels develop potent orthopoxviral-specific IgG antibody responses.

**Figure 2.**
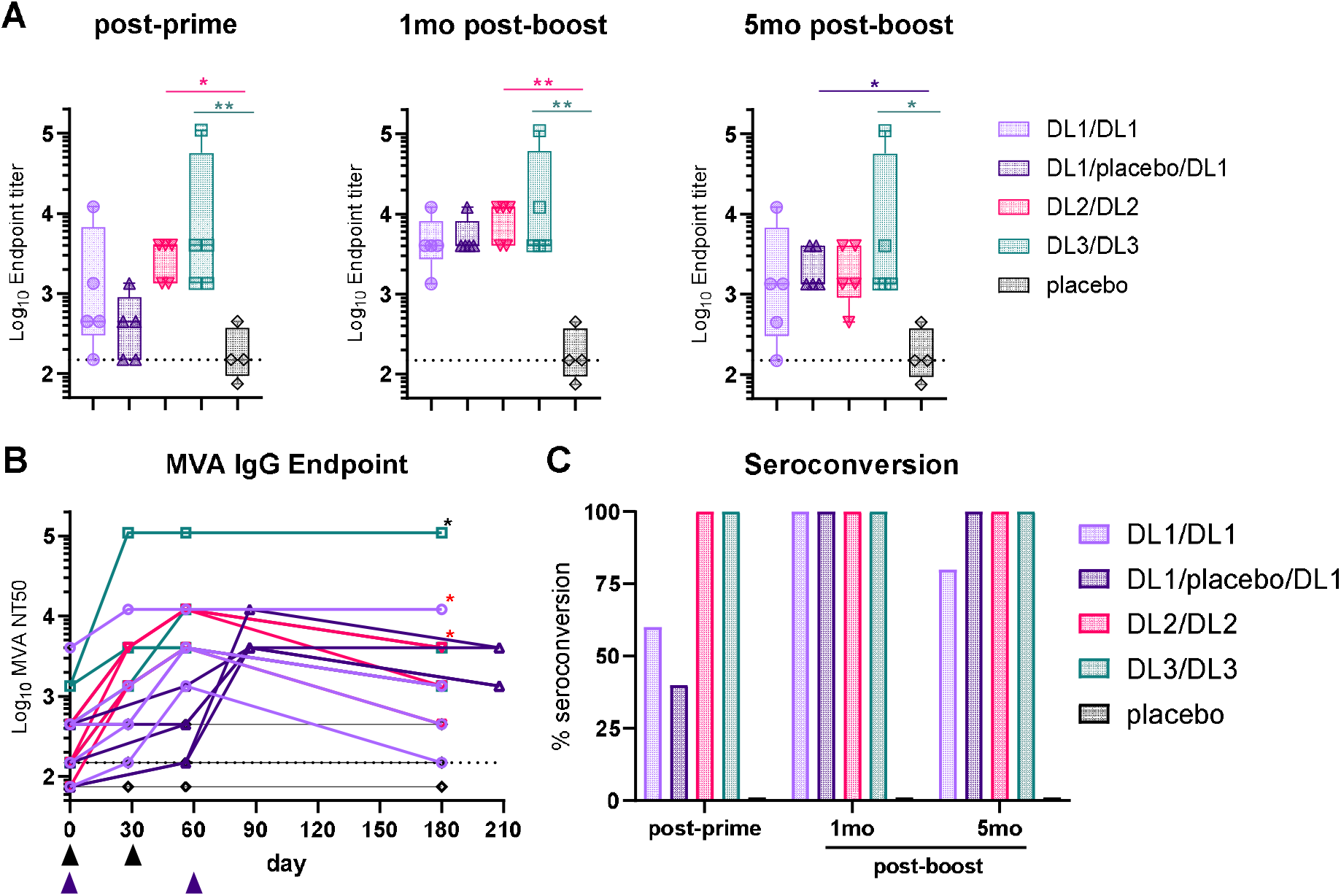
MVA-specific binding IgG in COH04S1 vaccinees. **A-B**. Binding antibodies. MVA-specific IgG endpoint titers were measured by ELISA in subjects before vaccination, post-prime vaccination, and at one- and five-months post-booster vaccination with COH04S1 at dose-level (DL) 1 (DL1/DL1 and DL1/placebo/DL1), DL2 (DL2/DL2), and DL3 (DL3/DL3). Subjects who received placebo vaccination were used as negative controls. Box plots in A extend from the 25^th^ to the 75^th^ percentiles, median values are shown as a line, whiskers extend from minimum to maximum values. Black triangles in B indicate time point of vaccination in DL1/DL1, DL2/DL2, and DL3/DL3 groups. Purple triangles indicate time of DL1 vaccinations in DL1/placebo/DL1 group. Red asterisks indicate subjects in DL1/DL1 and DL2/DL2 cohorts that were born before 1972. Black asterisk indicates the DL3 subject born in 1986 with suspected orthopoxvirus pre-existing immunity. Kruskal-Wallis test followed by Dunn’s multiple comparison test was used in A (*=p<0.05, **=p<0.01). **C**. Seroconversion rate. Shown is the percentage of seroconverted volunteers with MVA-specific NAb titers ≥3-fold above baseline at different time points post-vaccination with COH04S1.

### Orthopoxviral-specific neutralizing antibodies induced in COH04S1-vaccinated subjects

Similar to the observed MVA IgG responses in DL1-DL3 subjects, COH04S1-vaccinated subjects in all vaccine cohorts showed a strong increase in MVA-specific NAb titers, consistent with potent vaccine-induced orthopoxviral-specific immunity (Figure 3). Following prime vaccination, only a minor proportion of the DL1 subjects showed elevated MVA-specific NAb titers, whereas all DL2 and DL3 subjects showed an increase in MVA-specific NAb titers, confirming a dose-dependent vaccine effect after one dose (Figure 3A-B). While the DL1 cohorts showed a seroconversion rate of 0-60% after the first vaccination, DL2 and DL3 cohorts showed 100% seroconversion for MVA-specific NAb after the first dose (Figure 3C). All subjects of the different vaccine cohorts developed robust MVA-specific NAb titers at one month after the booster dose with NT50 titers ranging from 73 to >2,560 and a median NT50 of 303, resulting in 100% seroconversion in all vaccine cohorts. DL2 and DL3 subjects tended to have slightly higher NAb titers than DL1 vaccinated volunteers. Similar to the MVA IgG titers, MVA-specific NAb titers declined in all vaccine cohorts over five-months post-second vaccination, but they remained above baseline in most subjects, resulting in a median NT50 titer of 65 and comparable titers across vaccine groups. Only two volunteers in the DL2 and DL3 vaccine cohorts had undetectable MVA-specific NAb titers at five months after the second dose. Interestingly, the same two subjects with high baseline MVA binding IgG titers had pre-vaccination NAb titers approaching the NT50 detection limit of 20 and the highest MVA-specific NT50 across DL vaccine cohorts at five months post-boost, suggesting pre-existing orthopoxvirus-specific NAb responses in these vaccinees (Figure S3). Placebo-vaccinated volunteers had consistently undetectable MVA-specific NAb throughout the observation period (Figure 3). These results demonstrate that subjects vaccinated with COH04S1 at different dose levels develop robust and durable orthopoxvirus-specific NAb responses.

**Figure 3.**
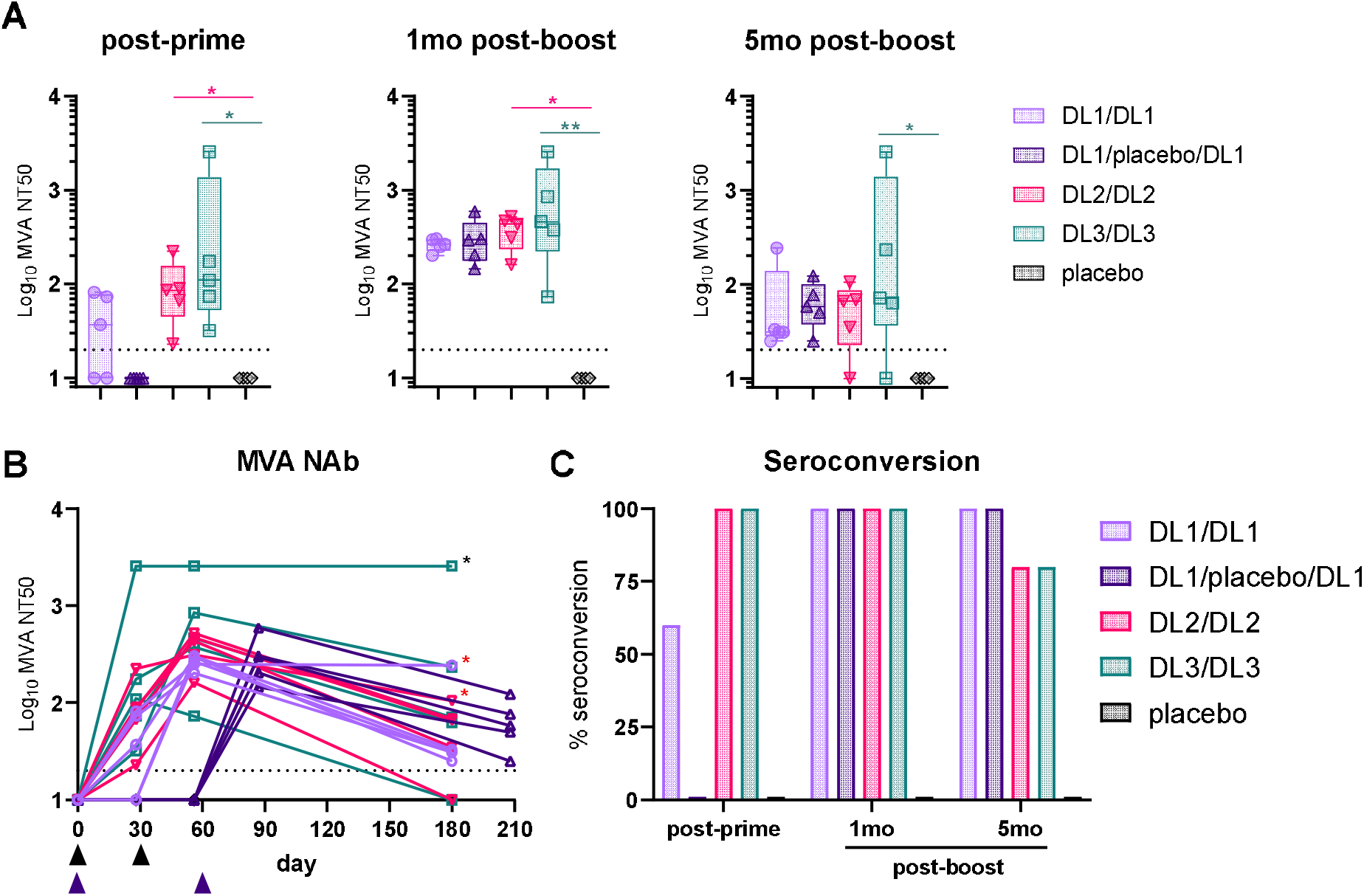
MVA-specific NAb responses in COH04S1 vaccinees. **A-B**. Neutralizing antibodies (NAb). MVA-specific NAb titers preventing 50% infection (NT50) were measured with a high-throughput neutralization assay. NAb were measured before vaccination, post-prime vaccination, and at one- and five-months post-booster vaccinations with COH04S1 at dose-level (DL) 1 (DL1/DL1 and DL1/placebo/DL1), DL2 (DL2/DL2), and DL3 (DL3/DL3). Subjects who received placebo vaccination were used as negative controls. Box plots in A extend from the 25^th^ to the 75^th^ percentiles, median values are shown as a line, whiskers extend from minimum to maximum values. Black triangles in B indicate time of vaccinations in DL1/DL1, DL2/DL2, and DL3/DL3 groups. Purple triangles indicate time of DL1 vaccinations in DL1/placebo/DL1 group. Red asterisks indicate subjects in DL1/DL1 and DL2/DL2 cohorts that were born <1972. Black asterisk indicates the DL3 subject born in 1986 with suspected orthopoxvirus pre-existing immunity. Kruskal-Wallis test followed by Dunn’s multiple comparison test was used in A (*=p<0.05, **=p<0.01). **C**. Seroconversion rate. Shown is the percentage of seroconverted volunteers with MVA-specific NAb titers above baseline at different time points post-vaccination with COH04S1.

### Orthopoxviral-specific cellular immunity induced in COH04S1-vaccinated subjects

Orthopoxvirus-specific T cells in COH04S1-vaccinated subjects were evaluated after the first and the second dose by assessing co-expression of IFNγ with CD107a or CD69 activation markers on MVA-stimulated T cells using flow cytometry (Figures S4-S5). CD107a marks cells capable of cytotoxic effector functions while CD69 is an early T cell activation marker that is transiently upregulated by activated T cells. Low levels of activated CD8^+^ and CD4^+^ T cells secreting IFNγ upon MVA stimulation were measured at baseline (Figures 4A and S6). COH04S1 vaccinees showed a significant increase in CD107^+^ and CD69^+^ IFNγ-secreting CD8^+^ and CD4^+^ T cells to maximal levels at one month after the first dose. After the second dose CD107^+^ and CD69^+^ IFNγ-secreting CD8^+^ and CD4^+^ T cells levels remained stable and significantly elevated levels of activated T cells were measured over five months post-boost.

**Figure 4.**
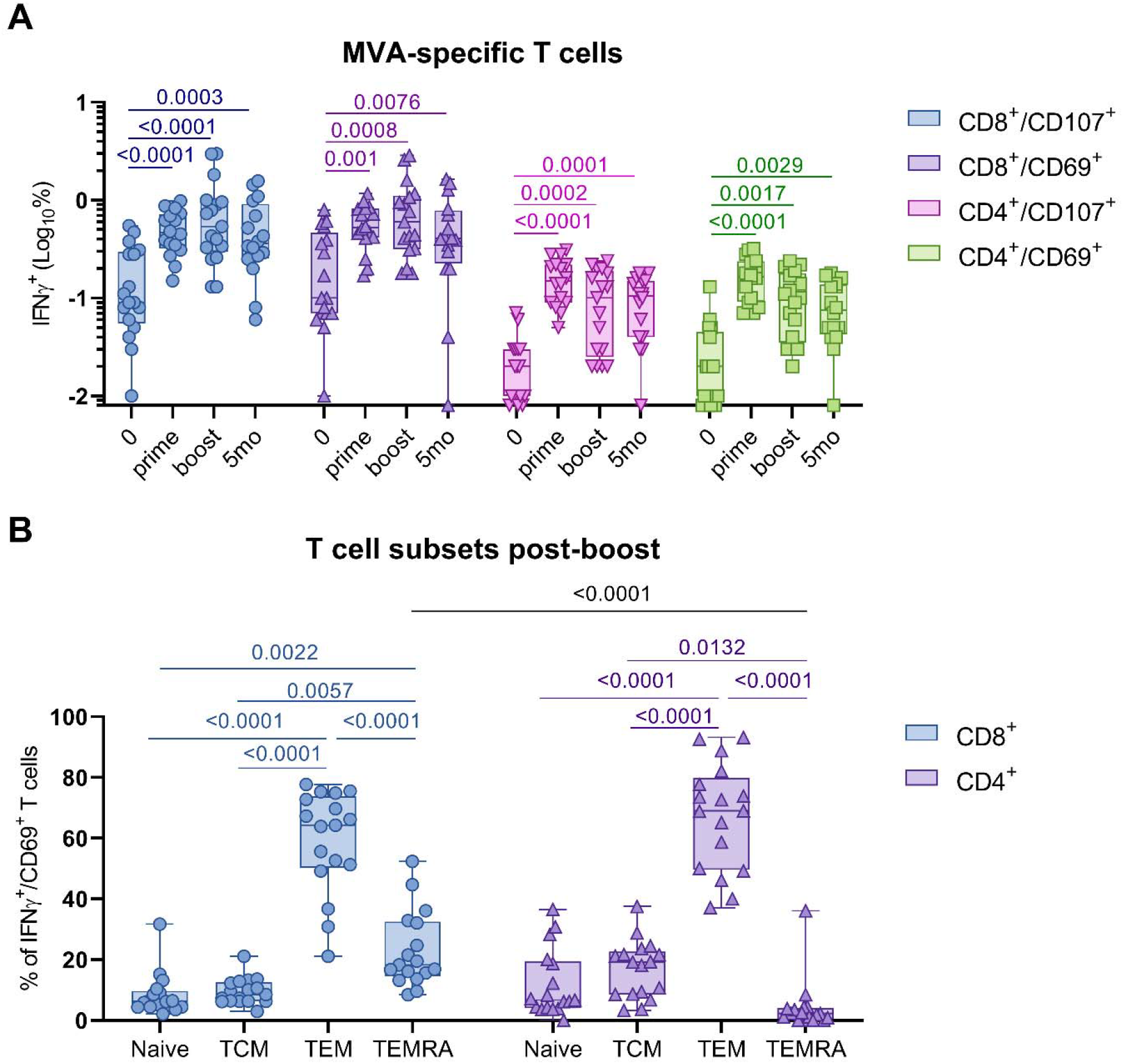
MVA-specific T cell responses in COH04S1 vaccinees. **A**. IFNγ^+^/CD107^+^ and γ /CD69 CD8 and CD4 T cell percentages were measured by cytofluorimetry in PBMC samples at baseline (0), at one-month post-prime (prime), and at one-month and five-months post-booster vaccinations with COH04S1 at all dose levels. Activated T cell percentages at baseline and after one or two vaccinations were compared using two-tailed Wilcoxon signed-rank test. **B**. Phenotypic analysis of antigen-specific T lymphocytes was performed using samples collected one-month post-second dose. Shown are percentages of naïve, central memory (T_CM_), effector memory (T_EM_), and terminally differentiated effector memory (T_EMRA_) T cells measured in IFNγ^+^/CD69^+^ CD8^+^ and CD4^+^ T cell populations. 2-way ANOVA followed by Sidak’s multiple comparison test was used to compare groups. P values are indicated in the figure. In A-B box plots extend from the 25^th^ to the 75^th^ percentiles, median values are shown as a line, whiskers extend from minimum to maximum values.

Similar levels of MVA-specific T cells were observed independently of dose level. In contrast, placebo subjects showed no or only low percentage of MVA-specific T cells (Figure S6).

Phenotypic analysis of activated MVA-specific T cell subsets revealed that both CD8^+^ and CD4^+^ T cell populations in COH04S1 vaccinees were mostly comprised of T effector memory (T_EM_) cells (Figure 4B and S7). Terminally differentiated T_EM_ cells (T_EMRA_) comprised about 20% of the CD8^+^ T cell population and were significantly higher than T_EMRA_ cells in the CD4^+^ population. Low percentages of naïve and central memory (T_CM_) T cells were measured in both CD8^+^ and CD4^+^ T cell populations. Comparable percentages of activated naïve/T_CM_/T_EM_/T_EMRA_ T cells were measured across the different DL vaccine cohorts (Figure S7). Finally, a similar phenotype distribution with predominance of T_EM/EMRA_ over T_CM_ was observed in the CD8^+^ and CD4^+^ T cell populations one-month after the first dose and one- and five-months post booster vaccination (Figure S7). These results demonstrate that at all tested dose levels vaccination with COH04S1 induces robust and durable orthopoxvirus-specific cellular responses with a predominant effector memory phenotype, whereby one vaccine dose is sufficient to obtain maximal induction of MVA-specific activated T cells.

### COH04S1 and sMVA elicit potent MPXV-specific antibodies in NHP and healthy adults

Next, we addressed whether COH04S1 and sMVA elicit MPXV-specific immune responses in vaccinated NHP by assessing binding antibodies to MPXV proteins H3 and A35, which are known targets of protective NAb. H3 is the intracellular mature virion (IMV) MPXV homologue of vaccinia H3L with which it shares 94% sequence, and it is the least conserved NAb target of the two members of the orthopoxvirus family^20^. Antibodies against H3L are likely a key contributor to protection against poxvirus infection and disease^20,21^. A35 is the more conserved MPXV homologue of vaccinia A33R protein which is associated with extracellular enveloped virions (EEV)^22,23^, the form of vaccinia infectious virus particle that is more resistant to NAb^24^.

After one sMVA or COH04S1 vaccine dose, NHP developed H3- and A35-specific IgG, with higher levels of H3- than A35-specific IgG (Figs. 5A-B, S8). At one-month after the second dose, increased MPXV-specific IgG titers were measured in both sMVA- and COH04S1-vaccinated NHP, indicating induction of comparable MPXV-specific antibodies by the two vaccines. No or low antibody responses to H3 and A35 proteins were measured in mock-vaccinated NHP.

**Figure 5.**
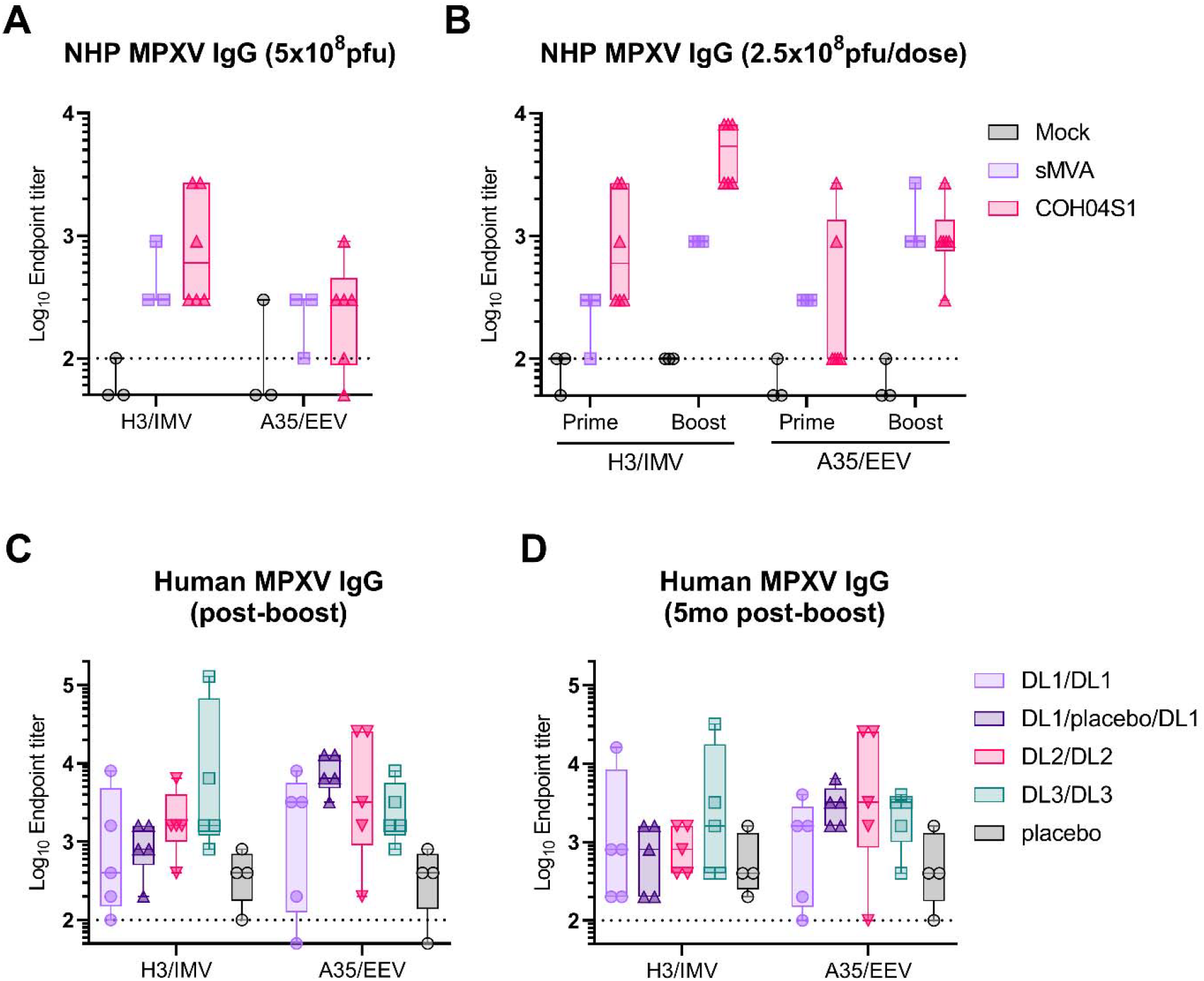
sMVA- and COH04S1-induced binding antibodies to MPXV antigens. **A-C**. MPXV-specific IgG endpoint titers to MPXV H3 and A35 proteins were measured by ELISA in NHP vaccinated once (A) or twice (B) with sMVA or COH04S1, and in healthy adults (C) one-month and (D) five-months post-booster vaccination with COH04S1 at dose-level (DL) 1 (DL1/DL1 and DL1/placebo/DL1), DL2 (DL2/DL2), and DL3 (DL3/DL3). Mock-vaccinated NHP and subjects who received placebo vaccination were used as negative controls. Dotted lines represent lower limit of detection. Box plots extend from the 25^th^ to the 75^th^ percentiles, median values are shown as a line, whiskers extend from minimum to maximum values. IMV= intracellular mature virions, EEV= extracellular enveloped virions.

In healthy subjects vaccinated with COH04S1, elevated H3- and A35-specific IgG were measured at one month after the second dose (Figs. 5C, S8). H3-specific IgG endpoint titers tended to be higher in subjects vaccinated at higher DL, while A35-specific IgG levels were more evenly distributed across DL vaccine cohorts. At five-months after the booster vaccination, MPXV-specific antibodies appear to decline, although elevated antibody titers to MPXV antigens A35 were consistently measured in COH04S1-vaccinees regardless of the DL used (Figs. 5D, S8). Interestingly, a strong correlation with vaccine-induced MVA-specific IgG was found for vaccine-induced H3-specific IgG but not for vaccine-induced A35-specific IgG (Fig. S9). These results demonstrate that MVA-based vaccines sMVA and COH04S1 induce MPXV-specific binding antibodies against antigens involved in the protection from MPXV infection.

## Discussion

In this report, we demonstrate that sMVA and multiantigen sMVA-based SARS-CoV-2 vaccine COH04S1 induce robust orthopoxvirus-specific immunity in NHP and healthy adults^18^. Our findings demonstrate that NHP vaccinated with one or two doses of either sMVA or COH04S1 develop potent othopoxvirus-specific antibody responses. Furthermore, we show that COH04S1-vaccinated volunteers receiving different vaccine doses develop robust MVA-specific humoral and cellular responses that remain detectable for up to six months post vaccination. Importantly, vaccination of NHP and healthy subjects with sMVA and COH04S1 induces elevated levels of binding antibodies recognizing MPXV proteins that are known target of protective NAb responses. These results suggest that sMVA and COH04S1 represent unique vaccine candidates that can be used as MPXV single-agent or MPXV/COVID-19 multi-agent vaccines as an effective countermeasure to mitigate the currently ongoing global MPXV health emergency.

MVA-specific binding and neutralizing antibodies developed in all volunteers after two vaccine doses resulting in 100% seroconversion. Consistent with a dose-escalation trial using a wild-type MVA^23^ (ACAM3000), we found that post-prime antibody titers were affected by the vaccine dose, with lower post-prime antibody titers in DL1 than in DL2/DL3 vaccinated subjects. However, following the second dose, COH04S1 was similarly immunogenic at all DL tested. On the contrary, no differences across DL were observed in magnitude and phenotype of MVA-specific activated T cells after one or two doses indicating that the lowest dose was sufficient to induce robust and durable cellular responses. This result is concordant with maximal induction of SARS-CoV-2 S- and N-specific T cells by COH04S1 at all DL tested as we have previously observed^18^.

As previously observed by others, early post-vaccine T cell response to orthopoxvirus antigens was largely comprised of CD8^+^ T cells^25,26^. Interestingly, COH04S1 induced higher MVA-specific CD8^+^ than CD4^+^ T cells, which is opposite to the previously observed SARS-CoV-2 S- and N-specific CD8^+^ and CD4^+^ T-cells induced by COH04S1^18^, indicating that different antigens can preferentially activate different T cell subtypes even during concomitant antigen stimulation. While it has been shown that T_EM_ are the predominant CD8^+^ subtype and T_CM_ the predominant CD4^+^ T cell subtypes in SARS-CoV-2 infected or vaccinated individuals^18,27,28^, we found that MVA-specific T_EM_ cells were the predominant subtype in both CD8^+^ and CD4^+^ T cell populations of COH04S1 vaccinees. Interestingly, T_EM_ cells, but not T_CM_ cells, have been demonstrated to protect against peripheral infection with vaccinia virus^29^, highlighting an important protective role of T_EM_ cells in orthopoxviral infections. Comprehensive studies of long-term immunity to vaccinia have shown that T_EM_ cells decay with time after antigen encounter while T_CM_ cells have a greater capacity to persist *in vivo*^30^. Whether the phenotype observed at six months post-vaccination in COH04S1-vaccinated subjects is maintained long-term or whether T_CM_ cells may overtake T_EM_ cells as the main MVA-specific T cell subpopulation can only be clarified in long-term studies.

Vaccination with sMVA-based vaccines stimulated roust binding antibodies to MPXV-specific NAb targets expressed on MPXV IMV and EEV forms^20,22^, demonstrating induction of broad cross-reactive orthopoxviral responses by sMVA-based vaccines. While IMV proteins are thought to play a predominant role in host-to-host transmission, EEV proteins are believed to be fundamental for dissemination within the host^22^. In previous studies, single IMV and EEV antigens were effective in partially protecting mice against lethal vaccinia challenge^31^, whereas a combination of four vaccinia IMV and EEV antigens was completely protective in mice and elicited MPXV-specific humoral responses in NHP^22^. Ultrapurified viral preparations have been shown to contain mostly IMV forms, which are released by cell lysis. Therefore, it is not surprising that we observed a highly significant correlation between MVA-specific IgG, which are measured using ultrapurified virus, and IgG to IMV MPXV antigen H3 but not to EEV MPXV antigen A35. Despite the absence of correlation, the presence of durable vaccine-induced EEV-specific antibodies demonstrated that the sMVA and COH04S1 vaccine products produced through ultrapurification were capable of inducing antibodies against the neutralization-resistant EEV form of the virus. This could be due to the presence of EEV proteins in the transitional intracellular enveloped virion (IEV) form^32^, which is released by cell lysis during the ultrapurification process together with IMV.

Smallpox and vaccinia immunity have been shown to be stable for decades after infection or vaccination and to decline only slowly over time^33,34^. Of the two volunteers born before 1972, the year of the end of the smallpox vaccination campaign, only one had an indication of low levels pre-existing poxviral immunity. However, both volunteers responded to two COH04S1 doses with higher-than-average and sustained MVA- and MPXV-specific humoral responses, suggesting that vaccination with COH04S1 successfully recalled low-to-undetectable vaccinia immunity acquired 50 years or more before, which resulted in more robust and durable responses than in naïve subjects. Interestingly, one DL3 subject born in 1986, and therefore not subjected to smallpox vaccination during childhood, showed low-level poxviral pre-existing humoral immunity at baseline. The same subject had a drastic increase in MVA-specific humoral response post-prime vaccination, and orthopoxvirus-specific binding antibodies and NAb at five months post-boost were exceptionally high. It is plausible that this subject was recently inoculated with vaccinia due to work exposure risk and that a combination of high COH04S1 dose with short time since smallpox vaccination contributed to the elevated orthopoxvirus-specific responses. Importantly, this subject and the two volunteers born before 1972 developed robust SARS-CoV-2 S- and N-specific humoral and cellular responses^18^, suggesting that vector-specific pre-existing immunity did not prevent induction of robust immunity to the SARS-CoV-2 antigens of COH04S1.

Definition of correlates of protection against smallpox and MPXV is complicated by the contrasting findings emerged in the past decades. Orthopoxvirus-specific NAb but not CD8^+^ T cells correlated with an attenuated Dryvax skin lesion or “take” at the inoculation site in one study^35^, and NAb were necessary and sufficient to protect monkeys against MPXV in another study^36^. On the other hand, a study in mice demonstrated an important protective role of T cell immunity in the absence of an antibody response^37^, and patients with defects in their T cell responses are known to be at risk to develop severe progressive vaccinia disease when vaccinated^38^. Consequently, it seems likely that a complex interplay of immunological factors contributes to the establishment of immunity to orthopoxviruses and these immunological correlates may vary based on species and viral strain. Therefore, it is encouraging that COH04S1 and sMVA vaccination induced a comprehensive orthopoxvirus-specific immunological response.

Major limitation of the study is the small number of subjects included in the study. Nonetheless, achievement of seroconversion and cellular responses in all subjects independent of vaccine dose indicates that vaccination with COH04S1 induces robust orthopoxvirus-specific immunity. Because of biosafety limitations, we have not used vaccinia or MPXV “live” viruses for assessment of vaccine immunogenicity. However, magnitude of MVA- and vaccinia-specific humoral immunity have been shown to be equivalent and we have tested induction of MPXV-specific immunity using MPXV antigens which are known targets of orthopoxvirus protective immunity^20,23,35^. A comparison of binding and NAb titers induced by COH04S1 and sMVA with titers measured in the WHO “International Standard for Anti-Smallpox Serum” 63/024^39^ was not possible since the product is currently not available in the NIBSC repository. Finally, to measure NAb, we utilized a high throughput neutralization assay based on the use of purified IMV. We have not evaluated vaccine-mediated neutralization of EEV although we measured EEV-specific IgG induced by COH04S1- and sMVA-vaccination using the MPXV EEV antigen A35.

Following the recent MPXV outbreak numerous countries have rushed ordering Jynneos for their nationals in need for a total of more than 3 million doses^40^. Additionally, the eventuality of a mass MPXV vaccination campaign in endemic African countries raises the issue of vaccine supply shortage and equitable distribution. Consequently, there is an urgency to replenish stockpiles around the world with safe and effective third generation smallpox/MPXV vaccines. The finding that both sMVA and COH04S1 induce robust and durable MVA- and MPXV-specific immunity represents the fundamental preliminary result for allowing COH04S1 and sMVA-based vaccines to be tested in non-inferiority clinical studies with immunological endpoints^9^ and challenge studies in non-human primates^41,42^ as vaccines against smallpox/MPXV.

## Methods

### Non-human primates

In life portion of NHP studies were carried out at Bioqual Inc. (Rockville, MD). The studies were conducted in compliance with local, state, and federal regulations and were approved by Bioqual and City of Hope Institutional Animal Care and Use Committees (IACUC). A total of 24 African green monkeys (Chlorocebus aethiops; 20 females and 4 males) from St. Kitts weighting 3–6□kg were randomized by weight and sex to vaccine and control groups. NHP were vaccinated twice four weeks apart with 2.5×10^8^ pfu of COH04S1 (n□=□6) or sMVA (n□=□3) diluted in PBS. Alternatively, NHP were vaccinated once with 5×10^8^ pfu of COH04S1 (n□=□6) or sMVA (n□=□3) diluted in PBS. Mock-vaccinated NHP immunized once (n□=□3) or twice (n□=□3) with PBS were used as additional controls. The evaluation of SARS-CoV-2 immunity following NHP-vaccination with COH04S1 has been previously described^16^.

### Human subjects

COH04S1 immunogenicity was investigated at City of Hope (COH) as part of a clinical protocol (IRB#20447) approved by an external Institutional Review Board (Advarra IRB). This open-label and randomized, placebo controlled, phase 1 clinical study is registered (NCT04639466). Among others, exclusion criteria included age<18 or >55, previous SARS-CoV-2 infection, BMI<18 or >35, underlying health conditions, and poxvirus vaccination within a six-months period. All subjects gave informed consent at enrollment. Out of the 51 subjects who received one or two doses of COH04S1, 5 subjects were selected from each dose group, for a total of 20 subjects, based on 2 doses regimen and availability of frozen PBMCs samples. Subjects received two doses of COH04S1 at days 0 and 28. Five subjects were vaccinated with dose level (DL) 1 (1×10^7^ pfu), five with DL2 (1×10^8^ pfu), and five with DL3 (2.5×10^8^ pfu). Additional five subjects received DL1 at day 0, placebo at day 28, and another DL1 at day 56. Four volunteers who received placebo at days 0 and 28 were included in the study. Two subjects - one in DL1/DL1 group and one in DL2/DL2 group-were born before 1972 and therefore may have had been previously vaccinated against smallpox. Study population is described on tables S1-S2. COH04S1-induced SARS-CoV-2 immunity in this population has been described before^18,19^.

### MVA and MPXV IgG Endpoint ELISA

MVA-specific binding antibodies were evaluated by ELISA. ELISA plates (3361, Corning) were coated overnight at 4°C with 1□µg/mL of MVA expressing Venus fluorescent marker (MVA-Venus)^13^, or with 10 µg/mL of A35 (40886-V08H), or H3L (40893-V08H1) MPXV antigens (SinoBiological) in PBS pH 7.4. Plates were washed 5X with wash buffer (0.1% Tween-20/PBS), then blocked with 250 µl/well of assay buffer (For NHP samples: 1% casein/PBS; for human samples: 0.5% casein/154mM NaCl/10mM Tris-HCl/0.1% Tween-20 [pH 7.6]/8% Normal goat serum) for 2 hours 37°C. After washing, 3-fold diluted heat-inactivated serum in blocking buffer was added to the plates. Plates were wrapped in foil and incubated 2 hours at 37°C after which plates were washed and 1:3,000 dilution of anti-human IgG HRP secondary antibody (BioRad 204005), or 1:10,000 anti-monkey IgG(H+L) HRP secondary antibody (Thermo Fisher PA1-84631) in assay buffer was added for 1 hour at room temperature. Plates were washed and developed with 1 Step TMB-Ultra (Thermo Fisher 34029). After 2-4 minutes the reaction was stopped with 1M H_2_SO_4_ and 450nm absorbance was immediately quantified on FilterMax F3 (Molecular Devices). Endpoint titers were calculated as the highest dilution to have an absorbance >0.100 nm. Seroconversion was defined as a three or more times increase in baseline titer.

### MVA neutralization assay

ARPE-19 cells were seeded in 96-well plates (1.5□×□10^4^ cells/well). The following day, 2-fold serial dilutions of serum starting from 1:10 were incubated for 2□h with MVA-Venus (multiplicity of infection [MOI]=2). The serum–virus mixture was added to the cells in duplicate wells and incubated for 24□h. After the 24□h incubation period, the cells were imaged using Leica DMi8 inverted microscope. Pictures from each well were processed using Image-Pro Premier (v9.2; Media Cybernetics) and fluorescent cells corresponding to infection events were counted. The neutralization titer for each dilution was calculated as follows: NT□=□[1-(fluorescent cells with immune sera/fluorescent cells without immune sera)]□×□100. The titers that gave 50% neutralization (NT50) were calculated by determining the linear slope of the graph plotting NT versus serum dilution by using the next higher and lower NT using Office Excel (v2019). Seroconversion was defined as an increase of two or more times the baseline titer^9^.

### Quantification of vaccine induced MVA-specific T cells

Peripheral blood mononuclear cells (PBMC) were isolated from fresh blood using Ficoll and counted using Luna-FL cell counter (Logos Biosystems). Frozen PBMCs were thawed, counted and 1×10^6^ PBMCs were stimulated with MVA-Venus (MOI=1) for 24 hours in a total volume of 200 µl of RPMI media with 5% of human serum in a 96 wells plate. Unstimulated cells and PHA (20 µg/ml) were used as negative and positive controls, respectively. Anti-CD107a-APC, Golgi Plug (Brefeldin A) and Golgi Stop (Monesin) were added 4 hours before staining. Cells were washed with PBS and stained 15 min at room temperature with Live and dead near IR, anti-CD3-FITC, anti-CD4-BV421, anti-CD8-BV605, anti-CD69-PE, anti-CCR7-PE/Dazzle 594 and anti-CD45-PerCP. After washing, cells were permeabilized with Fix/Perm (BD) for 20 minutes at 4°C. Cells were washed with Perm/Wash (BD) and intracellular stained with anti-IFNγ-PECy7 for 30 minutes at 4°C, washed and resuspend in FACS buffer until acquisition. Cells were acquired in Attune NxT cytometer (Thermofisher) and data was analyzed with Flow Jo X software following the gating strategy described in Figure S3. Only two out of five DL3 volunteers had available PBMCs samples for the analysis.

### Statistical analysis

Statistical analysis was performed using GraphPad Prism 8.3.0. Differences in humoral responses across groups were compared using Kruskal-Wallis test followed by Dunn’s multiple comparison test. T cell percentages at different time-points were compared using two-tailed Wilcoxon rank test. Differences in T cell subsets were evaluated using 2-way ANOVA followed by Sidak’s multiple comparison test. Pearson correlation coefficients and their p values were calculated for the correlative analysis.

## Data Availability

All data produced in the present work are contained in the manuscript

## Contributors

Study conceptualization: FC, FW, DJD. Study design: FC, FW. Immunological analysis: FC, SOF, ML. Manuscript writing: FC, FW, DJD. Clinical PI: JAZ. All authors contributed to and approved the final version of this manuscript.

## Data sharing

We support data sharing of the individual de-identified participant data that underlie the results reported in this article. Study protocols can be shared upon request by the corresponding author.

## Acknowledgements

The Authors would like to thank all the participants who volunteered in the study and all the investigators and study site personnel who assisted in the clinical trial completion. Funding was provided by the Carol Moss Foundation, donors Julie and Roger Baskes, Judd Malkin, Michael Sweig, and the City of Hope Integrated Drug Development Venture program. We acknowledge and thank Christoph Pittius and Yuriy Shostak (Research Business Development, City of Hope) for excellent project management. We thank Christina Ulloa (Department of Hematology & HCT, City of Hope) for excellent support of investigators and meeting coordination.

