## Supplement for "Synthetic modified vaccinia Ankara vaccines confer potent monkeypox immunity in non-human primates and healthy adults"

**Supplementary Material**

Table S1. Volunteers characteristics

| Characteristic | COH04S1,  N = 20 | DL1,  N = 10 | DL2,  N = 5 | DL3,  N = 5 | placebo,  N = 4 |
| --- | --- | --- | --- | --- | --- |
| Age^1^ | 36 (22, 54) | 39 (22, 50) | 40 (28, 54) | 32 (26, 41) | 31 (24, 41) |
| Gender |  |  |  |  |  |
| Female^2^ | 12 (60%) | 6 (60%) | 3 (60%) | 3 (60%) | 2 (50%) |
| Male^2^ | 8 (40%) | 4 (40%) | 2 (40%) | 2 (40%) | 2 (50%) |
| ^1^Median (Range); ^2^n (%). DL= dose level | | | | | |

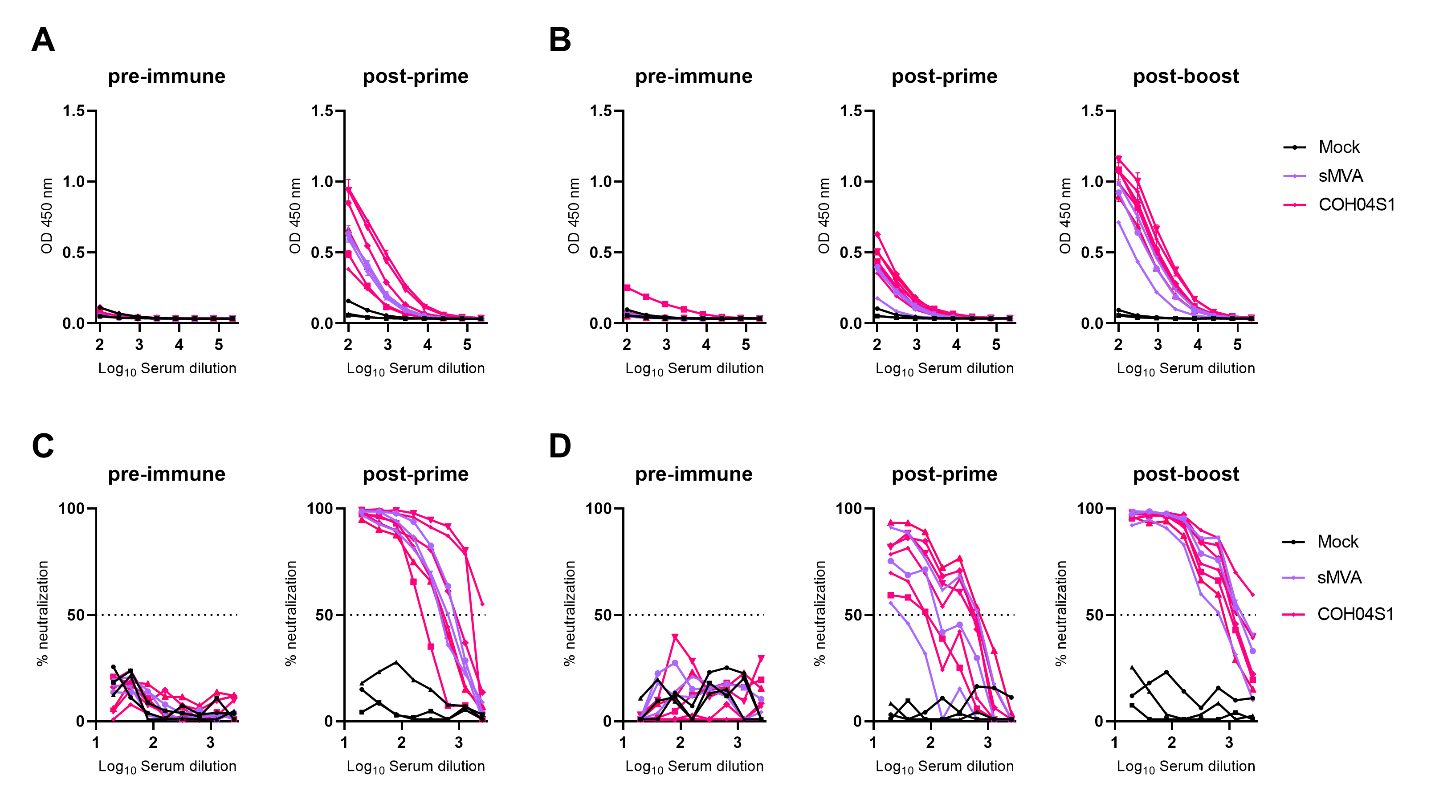

**Figure S1. Related to Figure 1. MVA-specific humoral response in sMVA- and COH04S1-vaccinated NHP.** NHP were vaccinated once with 5x10^8^ pfu (A, C) or two-times vaccinated with 2.5x10^8^ pfu (B, D) of sMVA (n=3) or COH04S1 (n=6). Mock-vaccinated NHP were used as controls (n=3). **A-B.** MVA-specific IgG endpoint titers were measured by ELISA at baseline, one month after the first dose, and one month after the second dose (in B). Shown are absorbance values (OD) measured at 450 nm. **C-D.** MVA-specific NAb titers. NAb specific for MVA were measured by microneutralization assay at baseline, one month after the first dose, and one month after the second dose (in D). Dotted lines represent 50% neutralization used to derive NT50.

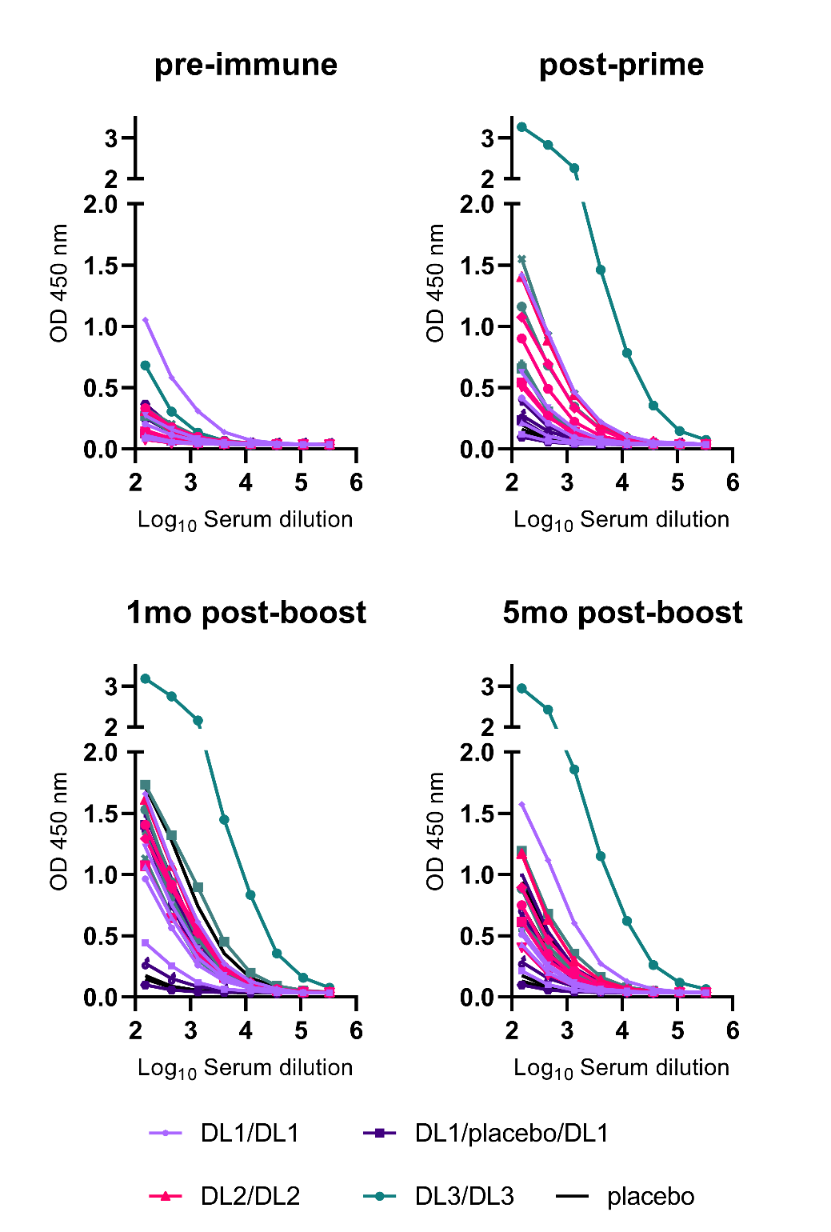

**Figure S2. Related to Figure 2. MVA-specific binding IgG in COH04S1 vaccinees.** MVA-specific binding IgG were measured by ELISA in healthy subjects before vaccination, post-prime vaccination, and at one- and five-months post-booster vaccination with COH04S1 at dose-level (DL) 1 (DL1/DL1 and DL1/placebo/DL1), DL2 (DL2/DL2), and DL3 (DL3/DL3). Subjects who received placebo vaccination were used as negative controls. Shown are the absorbance readings (OD) at 450 nm using serial dilutions of the serum.

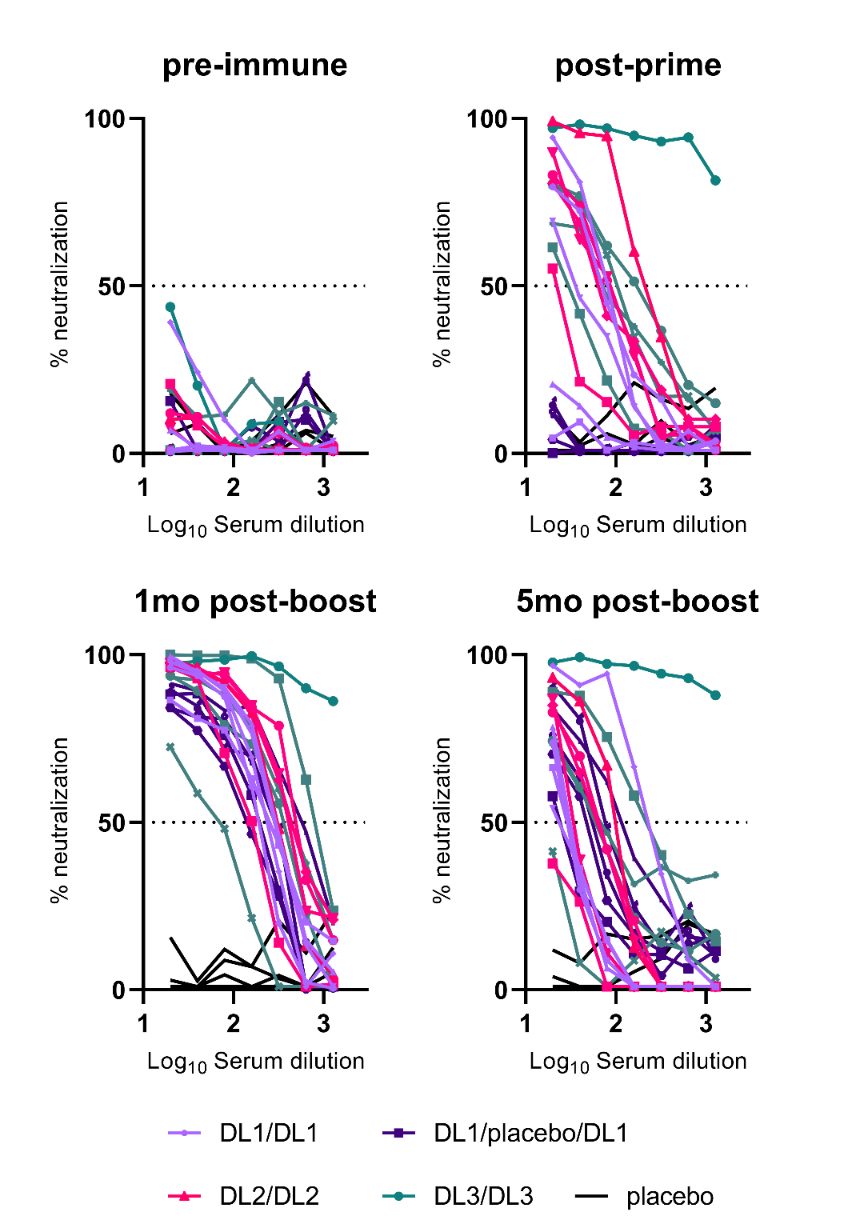

**Figure S3. Related to Figure 3. MVA-specific NAb titers in COH04S1 vaccinees.** MVA-specific NAb were measured using a high-thruput neutralization assay in serum samples of subjects before vaccination, post-prime vaccination, and at one and five months post-booster vaccinations with COH04S1 at dose-level (DL) 1 (DL1/DL1 and DL1/placebo/DL1), DL2 (DL2/DL2), and DL3 (DL3/DL3). Subjects who received placebo vaccination were used as negative controls. Shown are percentages of neutralization using serial dilutions of serum. Dotted lines mark the 50% neutralization used to derive NT50 titers.

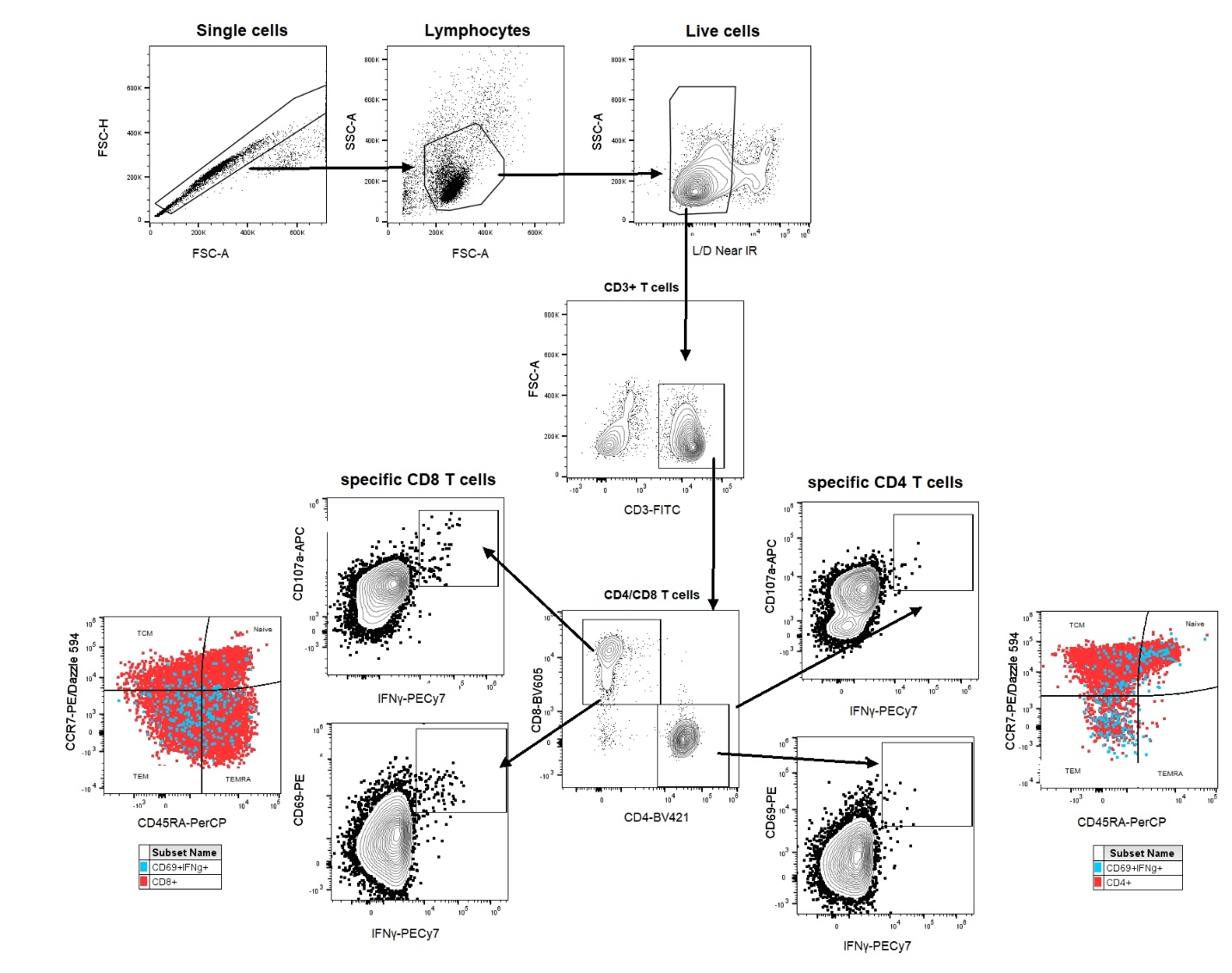

**Figure S4. Related to Figure 4. MVA-specific T cell response in COH04S1 vaccinees.** Gating strategy. Shown is the gating strategy used to identify MVA-specific T cells. Gating was performed on single cells>lymphocytes>live cells>CD3^+^ cells> CD8^+^ or CD4^+^ cells> CD107a^+^/IFNγ^+^ or CD69^+^/IFNγ^+^ cells. CD69^+^/IFNγ^+^ double positive cells were used to identify CD8^+^ and CD4^+^ T cell memory subsets. Naïve cells were identified as CCR7^+^/CD45RA^+^; central memory (T_CM_) cells were identified as CCR7^+^/CD45RA^-^; effector memory (T_EM_) cells were identified as CCR7^-^/CD45RA^-^; and terminally differentiated effector memory (T_EMRA_) cells were identified as CCR7^-^/CD45RA^+^.

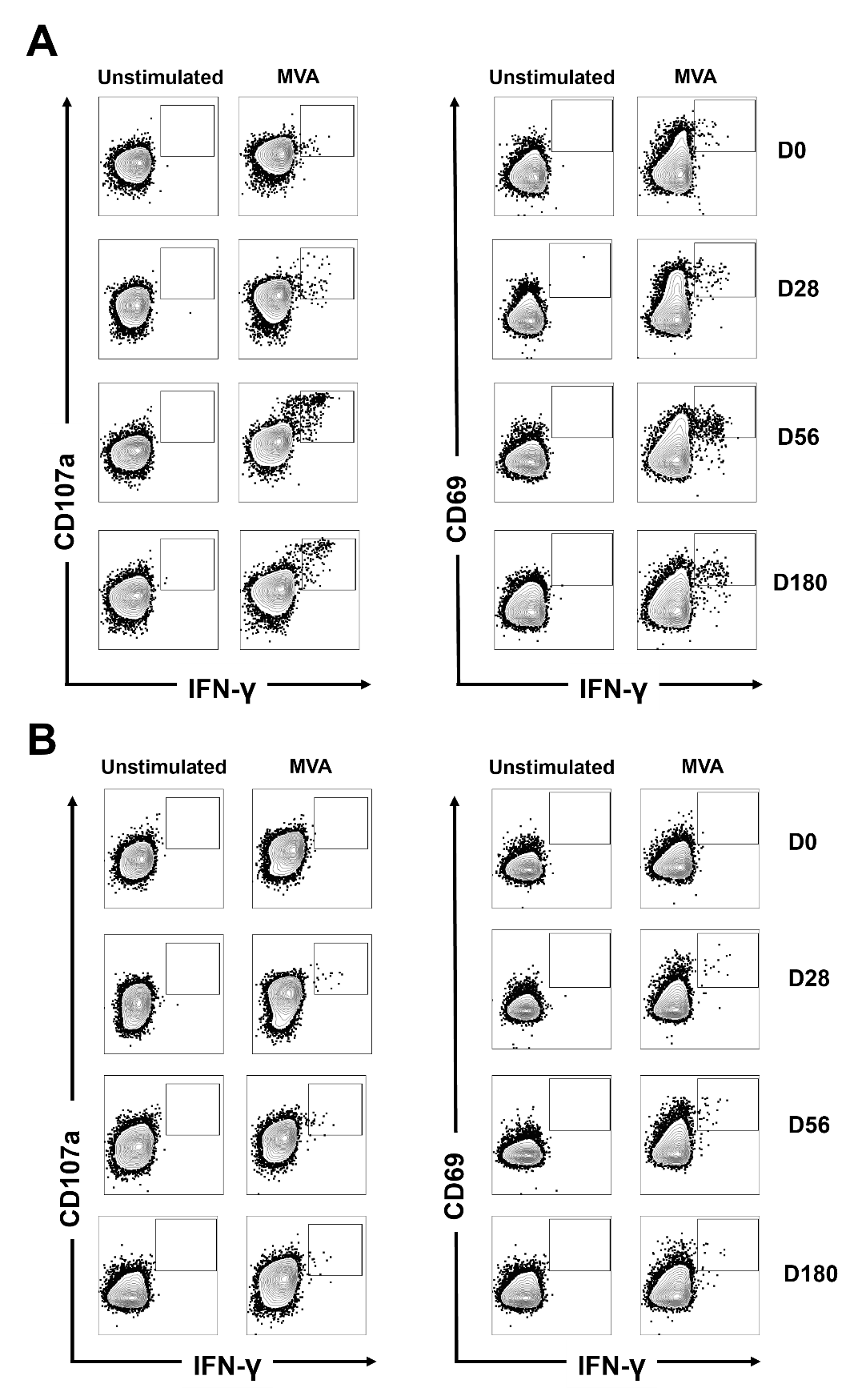

**Figure S5. Related to Figure 4. MVA-specific T cell response in COH04S1 vaccinees.** Example of T cell analysis. Shown is an example of gating of double positive CD107a^+^/CD69^+^ and IFNγ^+^ CD8^+^ (A) or CD4^+^ (B) T cells measured in unstimulated or MVA-stimulated PBMC samples of a volunteer (DL1) before and after vaccination with COH04S1.

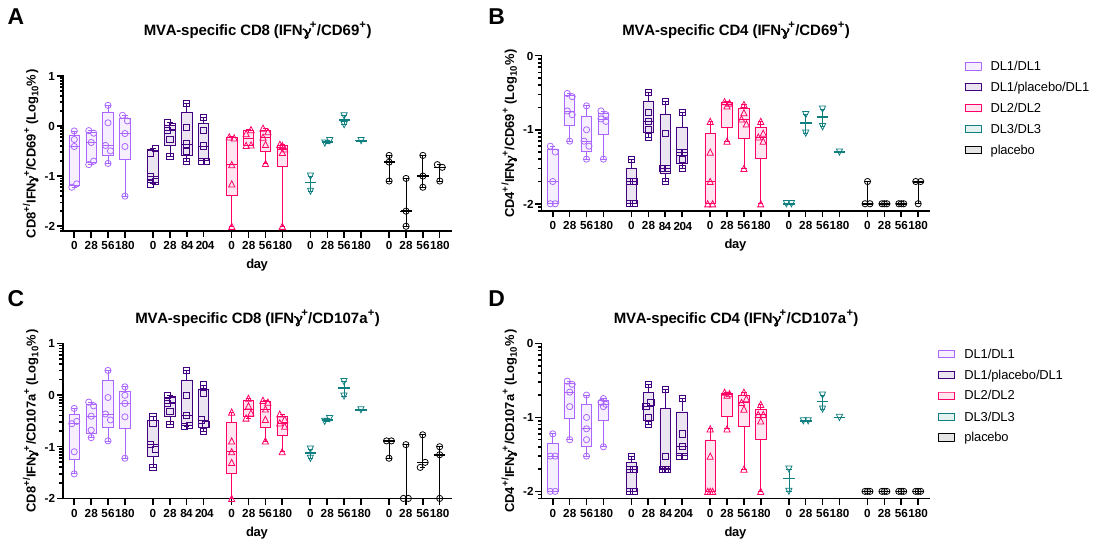

**Figure S6. Related to Figure 4. MVA-specific T cell response in COH04S1 vaccinees.** Shown are activated MVA-specific T cells after vaccination with COH04S1. Percentages of MVA-specific IFNγ^+^/CD69^+^ CD8^+^ (A) and CD4^+^ (B) T cells, and MVA-specific IFNγ^+^/CD107^+^ CD8^+^ (C) and CD4^+^ (D) T cells were measured in PBMC samples by cytofluorimetry at baseline, one-month after the first vaccination, and one- and five-months post-booster vaccinations with COH04S1 at dose-level (DL) 1 (DL1/DL1 and DL1/placebo/DL1), DL2 (DL2/DL2), and DL3 (DL3/DL3). Subjects who received placebo vaccination were used as negative controls. Only two DL3 volunteers had available PBMC samples for the analysis.

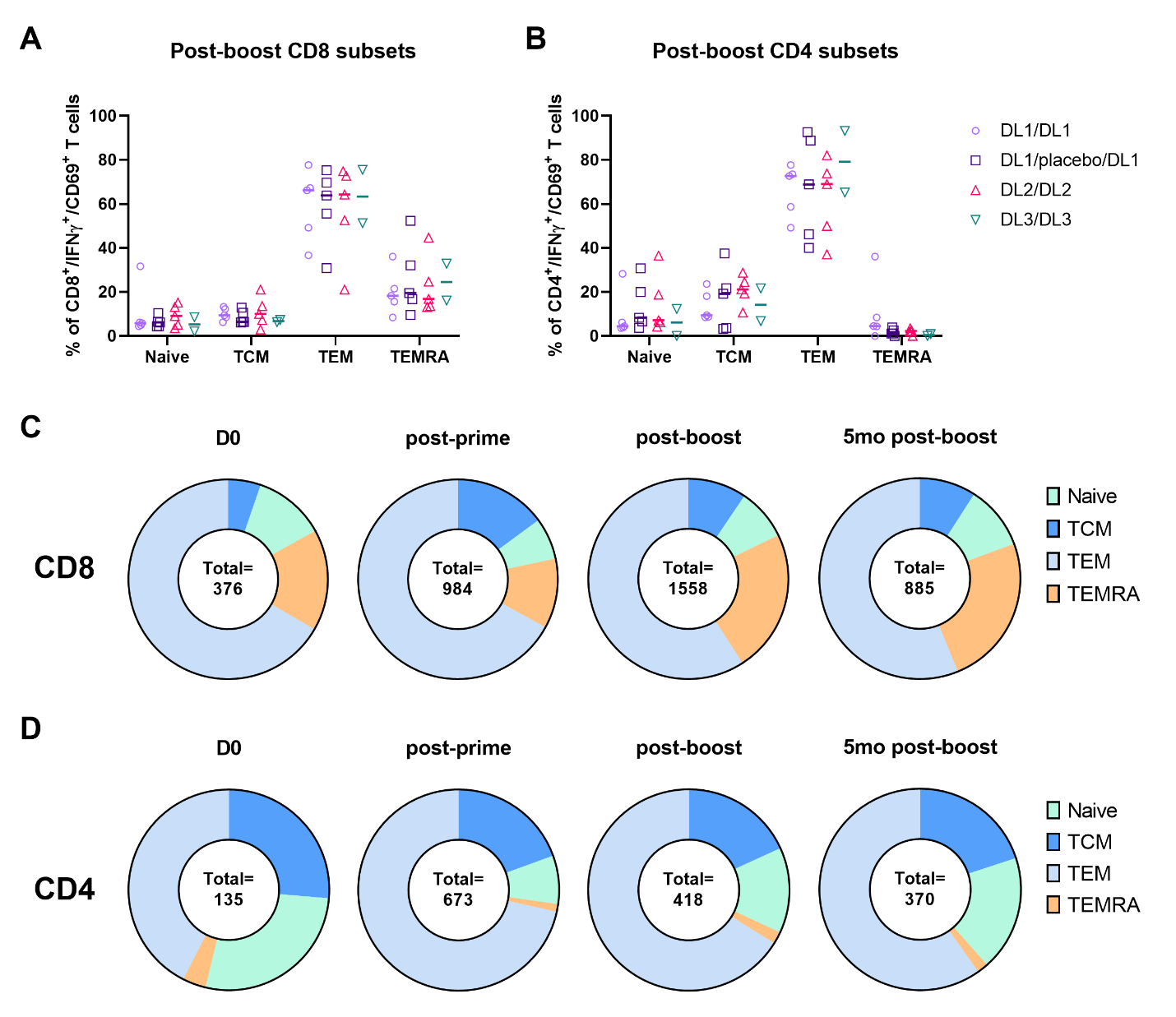

**Figure S7. Related to Figure 4. MVA-specific T cell response in COH04S1 vaccinees.** Shown are activated T cell memory subtypes after vaccination with COH04S1. **A-B.** Phenotypic analysis of antigen-specific T lymphocytes was performed using samples collected one month after the second dose of COH04S1 at dose-level (DL) 1 (DL1/DL1 and DL1/placebo/DL1), DL2 (DL2/DL2), and DL3 (DL3/DL3). Shown are percentages of naïve, central memory (TCM), effector memory (TEM), and terminally differentiated effector memory (TEMRA) T cells measured in IFNγ+/CD69+ CD8^+^ (A) and CD4^+^ (B) T cell populations. Median values are indicated with lines. **C-D.** Relative contribution of activated CD8^+^ (C) and CD4^+^ (D) T cell memory subtypes in subjects vaccinated with COH04S1 at baseline, one-month after the first vaccination, and one- and five-months post-second dose. Total indicates the average number of IFNγ+/CD69+ CD8+ or CD4+ T cells/100 μl of blood.

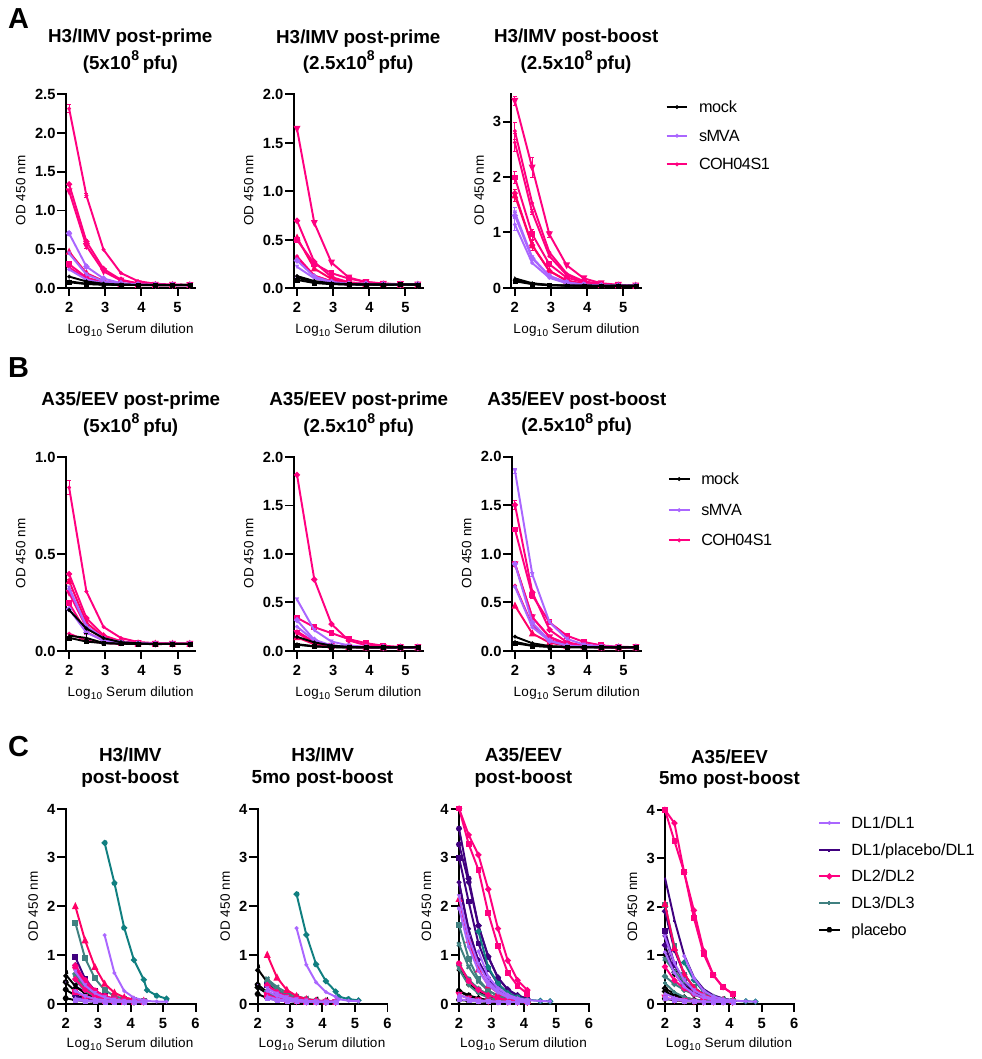

**Figure S8. Related to Figure 5. MPXV-specific binding IgG in NHP and healthy subjects vaccinated with sMVA and COH04S1.** MPXV-specific IgG endpoint titers to MPXV H3 and A35 proteins were measured by ELISA in NHP vaccinated once and twice with sMVA or COH04S1 (A-B), and in healthy adults (C) one-month and five-months after-booster vaccination with COH04S1 at dose-level (DL) 1 (DL1/DL1 and DL1/placebo/DL1), DL2 (DL2/DL2), and DL3 (DL3/DL3). Mock-vaccinated NHP and subjects who received placebo vaccination were used as negative controls. Shown are absorbance (OD) values at measured at 450 nm.

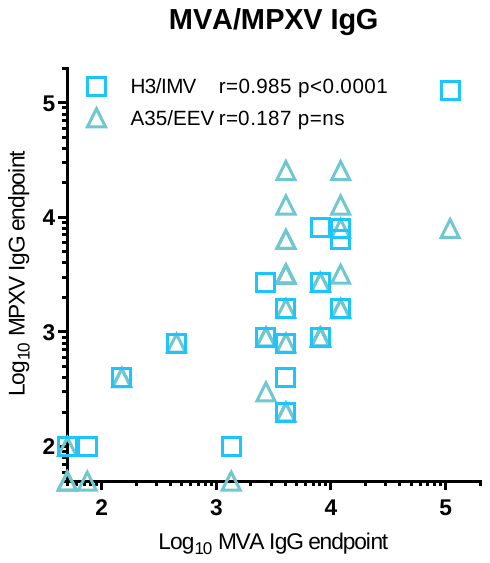

**Figure S9. Related to Figure 5. Correlative analysis of MVA- and MPXV-specific binding antibodies.** MVA-specific endpoint titers measured in NHP and human serum samples were correlated to MPXV-specific endpoint titers measured in the same serum samples using H3 and A35 proteins. Shown are Pearson’s correlation coefficients (r) and their two-tailed significance (p).
